# Measuring Quality-of-Care in Treatment of Children with Attention-Deficit/Hyperactivity Disorder: A Novel Application of Natural Language Processing

**DOI:** 10.1101/2023.06.12.23291071

**Authors:** Malvika Pillai, Jose Posada, Rebecca M. Gardner, Tina Hernandez-Boussard, Yair Bannett

**Author notes:** **Address correspondence to:** Yair Bannett, Developmental-Behavioral Pediatrics, Stanford University School of Medicine, 3145 Porter Drive, Palo Alto, CA, 94304.

## Abstract

**Objective:** To develop and validate a novel application of natural language processing (NLP) techniques to measure pediatrician adherence to evidence-based guidelines in the treatment of young children with attention-deficit/hyperactivity disorder (ADHD).

**Materials and Methods:** We extracted structured and free text data from electronic health records of all office visits (2015-2019) of children aged 4-6 years seen in a community-based primary healthcare network in California, who had ≥1 visits with an ICD-10 diagnosis of ADHD. Two pediatricians manually annotated clinical notes of the first ADHD visit for 423 patients. Inter-annotator agreement was assessed for recommendation for first-line behavioral treatment; Disagreements were reconciled. The BioClinical Bidirectional Encoder Representations from Transformers (BioClinical-BERT) was used to identify mentions of behavioral treatment recommendations using a 70/30 train/test split. Following an error analysis and threshold selection, we completed external (temporal) validation by deploying the model on 1,020 unannotated notes representing other ADHD visits and well-care visits; all positively classified notes and 5% of negatively classified notes were annotated.

**Results:** Of 423 included patients, 313 (74%) were male, 268 (63%) were privately insured; 138 (33%) were white; 61 (14 %) were Hispanic. The BERT model of first ADHD visits achieved F1=0.78, precision=0.84, and recall=0.72. Following threshold selection, temporal validation on notes from other visits achieved F1=0.80, recall=0.92 and precision=0.7.

**Conclusion:** Deploying a machine learning algorithm on a large and variable set of clinical notes accurately captured pediatrician adherence to guidelines in treatment of children with ADHD. This approach can be used to measure quality-of-care at scale and improve clinical care for various chronic medical conditions.

## INTRODUCTION

Attention-Deficit/Hyperactivity Disorder (ADHD) is the most common child neurobehavioral disorder, estimated to affect 8-10% of US children.^1,2^ Most children with ADHD are diagnosed in the preschool or elementary school period of life, and are at high risk for academic failure.^3-6^ The primary care pediatrician is most often the professional who manages the treatment of ADHD in young children.^7,8^ The American Academy of Pediatrics (AAP) distributed and updated evidence-based clinical practice guidelines for primary care management of ADHD in 2001, 2011, and 2019.^9-11^ For 4-5 year old preschoolers with ADHD or ADHD symptoms, guidelines emphasize parent training in behavior management (PTBM) as the first-line treatment based on stronger evidence for PTBM than for medications for treatment of preschoolers with or at-risk for ADHD.^5^ For 6-11-year-old school aged children with ADHD, guidelines recommend combining PTBM along with medications based on the evidence suggesting a combined therapy in this age group is superior to either therapy alone in improving child and family functioning.^11-13^

The National Institute for Children’s Health Quality has recognized that quality measures for child mental health disorders, including Attention-Deficit/Hyperactivity Disorder (ADHD), are lacking.^14,15^ The current national quality measures used for assessment of ADHD care include the Healthcare Effectiveness Data and Information Set (HEDIS) measures that capture the timing of follow up care for children prescribed with ADHD medications.^16^ These claims-based measures are readily available and easily calculated, but they only address a narrow aspect of care in a subset of patients with ADHD. Furthermore, these measures have received a “poor” or “not-rated” strength of evidence grade.^15^ Recognizing the significant limitations of the HEDIS measures, many health care organizations supplement these crude measures with labor-intensive and expensive manual chart review of the electronic health record (EHR) of a sample of ADHD patients per clinician.^17,18^

The few EHR-based studies that objectively assessed ADHD care by primary care providers (PCPs) – through manual review of medical records – found quality gaps in low adherence to several components of published guidelines, including low rates of recommendations for PTBM, use of validated rating scales, and appropriate follow up.^19-21^ The lack of scalable access to essential components of clinical care that are documented as free text in the EHR is the major barrier preventing development of process-of-care quality indicators for primary care management of ADHD that rely on evidence-based clinical practice guidelines. Machine learning techniques of natural language processing (NLP) offer a unique opportunity to analyze at a large scale free-text information in the EHR that captures the entire set of recommendations given to families.^22,23^

In this study, we utilized BioClinical-BERT (BioClinical Bidirectional Encoder Representations from Transformers), a machine learning-based classification models to analyze unstructured (free-text) data from the EHR of a large community-based pediatrics primary care network to assess the rate of PCP recommendations and counseling regarding PTBM in children aged 4-6 years who presented with ADHD or related symptoms.

## METHODS

We present the study in accordance with the MINimum Information for Medical AI Reporting (MINIMAR) framework.^24^ This study was approved by the Stanford University School of Medicine institutional review board.

### Setting and Population

Packard Children’s Health Alliance (PCHA) is a community-based pediatric healthcare network in the San Francisco Bay Area, affiliated with Stanford Children’s Health and Lucile Packard Children’s Hospital. PCHA has 24 pediatrics primary care offices, grouped into 10 practices. We confirmed that visit diagnosis codes are clinician-entered in all practices.

### Data Sources

We conducted a retrospective review of EHRs for a cohort of all pediatric patients seen by PCHA PCPs from October 1, 2015 (date coinciding with adoption of ICD-10 codes) to December 31, 2019. We extracted de-identified structured and free-text data from all office encounters. We reviewed patient records of children aged 4-6 years who had at least 2 visits during the examined period, and who had at least one visit in the study period with a disorder (ADHD) <u>or</u> symptom-level diagnosis code (e.g., hyperactivity, distractibility), as done in our previous study (n=423).^25^ We excluded patients with a diagnosis of autism in the study period. **Figure 1** shows the study workflow.

**Figure 1.**
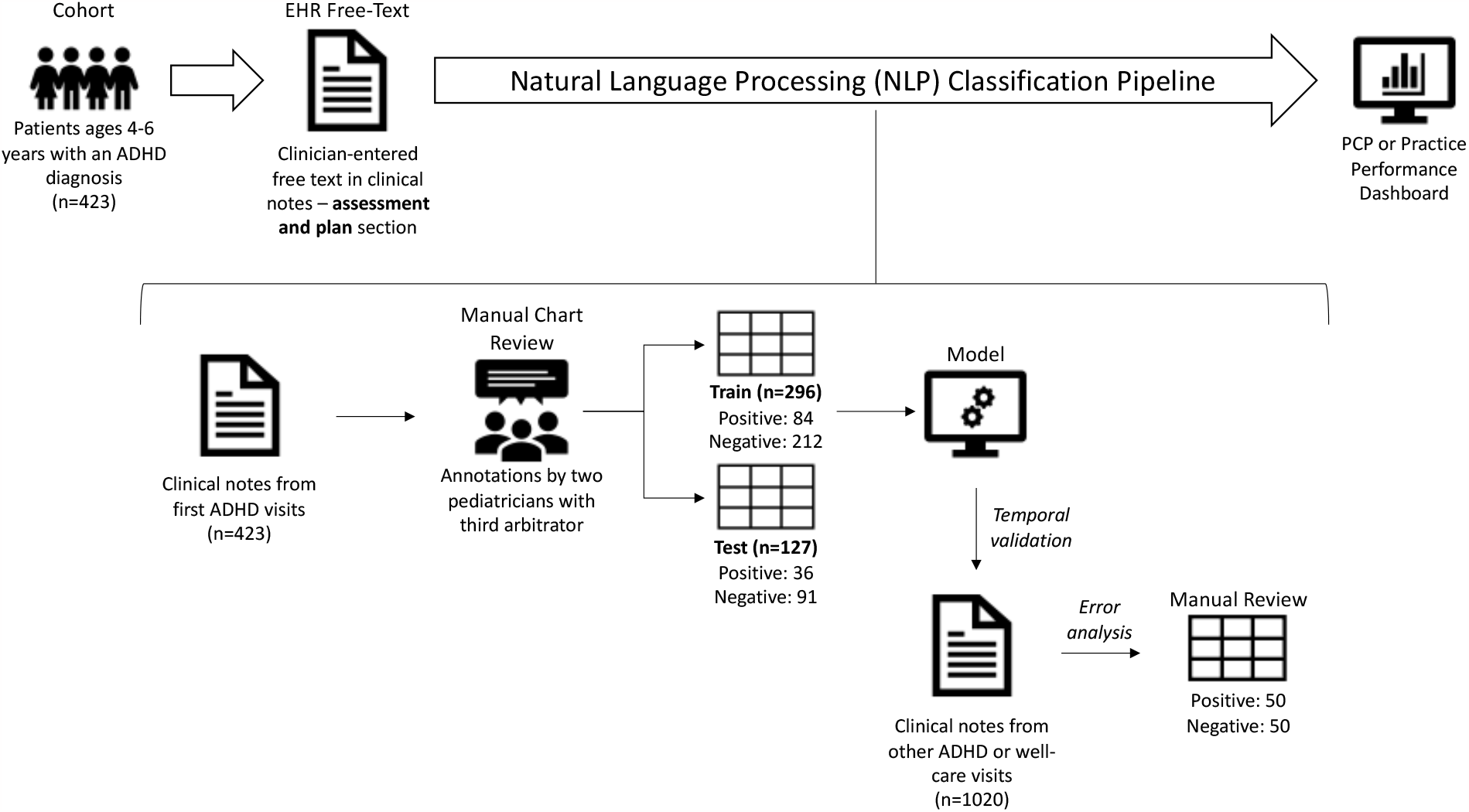
Study workflow

### Manual chart review and annotation

The primary outcome of interest was rate of PCP recommendation of Parent Training in Behavioral Management (PTBM) as part of treatment recommendations in the first visit with an ADHD diagnosis. PTBM recommendations included referral to PTBM and/or counseling the family regarding behavioral management (e.g., providing a handout). To create a “gold standard” for mention of PTBM by PCPs, we developed annotation guidelines to be used by two pediatricians (YB, ST) who performed independent chart review and annotation of the first visit with an ADHD diagnosis for each child in the study cohort.^26^ Inter-annotator agreement was assessed for mention of PTBM. In step 1, document-level annotations of PTBM mentions were compared between the two annotators and disagreements were used to refine the annotation guidelines. In step 2, annotation guidelines were further refined using the same method. In step 3, the annotators completed the annotation of all notes, and all disagreements (13%) were reviewed and discussed. In cases an agreement could not be reached, a 3^rd^ pediatrician (L.H.) was used to reach a decision.

### Data preparation

We annotated the 423 first ADHD visit notes (i.e., the first visit with a visit diagnosis of ADHD), which we considered to have the highest likelihood of PTBM recommendations. We identified an additional 1020 other notes from all subsequent (follow-up) ADHD visits and all well-care visits between the ages of 4-6 years, which we thought may have included recommendations for PTBM. The annotated set of first ADHD visit notes for the cohort was randomly subset into train (n=296) and test (n=127) sets with a 70-30 split. The train set was used for model development and hyperparameter tuning while the test set was set aside to evaluate model performance on first visits. Temporal validation was conducted by deploying the model on all other patient notes from ADHD or well-care visits (n=1020), which were unannotated.

Notes pre-processing was divided into data cleaning and extraction. All notes were processed through a basic cleaning pipeline that included stripping punctuation, extra whitespace, and digits. A pipeline for extracting sections of interest from the notes was created. The notes were organized in the SOAP (Subjective/Objective/Assessment/Plan) structure. To focus on treatment recommendations by clinicians, the chart review was limited to the Assessment and Plan sections where clinicians are expected to document their impressions, clinical reasoning, and treatment recommendations.

### NLP model development

We developed a binary classification pipeline based on various language models to classify notes as containing recommendations for PTBM or not. Six transformer models were used for this task: Bidirectional Encoder Representations from Transformers (BERT) uncased, RoBERTa, XLNet, ClinicalBERT, and BioClinicalBERT. BERT uncased is a variation of the original BERT model that is only trained on lowercase text from general purpose corpora. RoBERTa (Robustly Optimized BERT Approach) was trained on diverse general-purpose corpora. XLNet (eXtreme Language understanding NETwork) and is based on BERT and RoBERTa. It differs from BERT models in that it learns dependencies between words in different contexts by predicting all permutations of the words in a sequence. BioClinicalBERT was trained on MIMIC III notes but was also initialized from BioBERT, which was trained on biomedical corpora.

For each transformer model, a classification layer was incorporated that generated softmax values for each set of scores. Hyperparameter tuning was done on the training set, and performance was evaluated on the test set using various metrics including F1-score, accuracy, precision, recall, and area under the receiver operating characteristic curve (AUROC). Because of low prevalence of positive cases in our dataset, we also evaluated the best performing model with the area under the precision-recall curve (AUPRC), which is a metric that is less sensitive to imbalanced classes. Model thresholds were selected on the precision-recall curve to maximize recall and minimize the false negative rate on the train set. As this work aims to assess clinician adherence to guidelines, we intended to avoid wrongly penalizing clinicians by minimizing the false negative rate. The top-performing model on the test set (n=127) was chosen for temporal validation. The code is available via Github at https://github.com/ybannett/NLP_ADHD.

### Temporal validation and error analysis

The best model was deployed on all other ADHD or well-care visit notes (n=1020). All notes classified as positive (n=50) and a random subset of 50 notes (5%) classified as negative were manually reviewed. The misclassified notes were analyzed to understand model errors and potential reasoning behind misclassifications.

## RESULTS

### Cohort characteristics

This study included 423 patients aged 4-6 years with an ADHD diagnosis. Based on manual chart review of all first visits with an ADHD diagnosis, PTBM was recommended for 30.5% of patients (n=129) at their first visit. Cohort characteristics by PTBM recommendation (yes/no) are displayed in **Table 1**. Of 423 patients, the average patient age at first ADHD visit was 5.4 years with 74% being male. The cohort primarily consisted of non-Hispanic white patients (n=138, 32.6%) and patients with unknown race/ethnicity (n=131, 31.2%). Most patients were privately insured (n=268, 63.4%); about a third were publicly insured (n=124, 29.3%). Within race/ethnicity subgroups, 33.3% (46/138) of non-Hispanic white patients received behavioral treatment recommendations compared to only 16.7% (4/24) of non-Hispanic black patients. When examining the cohort by insurance type, 32.5% (87/268) of patients with private insurance received behavioral treatment recommendations compared to only 28% (35/124) of patients with public insurance.

**Table 1.** Patient cohort characteristics (n=423)

| CHARACTERISTICS | N = 423 |  |  |  |  |
| --- | --- | --- | --- | --- | --- |
|  | Behavioral treatment recommended |  |  |  |  |
|  | No<br>(n = 294) |  | Yes<br>(n = 129) |  | Standardized<br>Mean<br>Difference<br>(SMD)<sup>a</sup> |
|  | n | % | n | % |  |
| **Race/ethnicity** |  |  |  |  | 0.274 |
| Non-Hispanic White | 92 | 31.3 | 46 | 35.7 |  |
| Non-Hispanic Asian | 23 | 7.8 | 14 | 10.9 |  |
| Non-Hispanic Black | 20 | 6.8 | 4 | 3.1 |  |
| Hispanic | 47 | 16.0 | 14 | 10.9 |  |
| Non-Hispanic Other | 19 | 6.5 | 12 | 9.3 |  |
| Unknown | 93 | 31.6 | 39 | 30.2 |  |
| **Average Age at First**<br>**ADHD Visit** | 5.48 |  | 5.22 |  | 0.396 |
| **Sex** |  |  |  |  | 0.114 |
| Male | 213 | 72.4 | 100 | 77.5 |  |
| Female | 81 | 27.4 | 29 | 22.5 |  |
| **Insurance** |  |  |  |  | 0.156 |
| **Private** | 181 | 61.6 | 87 | 67.4 |  |
| **Public** | 89 | 30.3 | 35 | 27.1 |  |
| **Military** | 23 | 7.8 | 7 | 5.4 |  |
| **Unknown** | 1 | 0.3 | 0 | 0 |  |

<sup>a</sup>Standardized Mean Difference values of 0.2, 0.5, and 0.8 correspond to small, moderate, and large differences, respectively.

### Classification results on test set

Five models were compared to classify clinical notes as including a PTBM recommendation or not. Models were trained using the training set (n=296), and performance was evaluated on the test set (n=127) (**Table 2**). The highest performing model was BioClinicalBERT with an F1-score of 0.78 followed by XLNet with an F1-score of 0.71.

**Table 2.** Model performance classifying clinical notes in the test set

| **Model** | **Precision** | **Recall** | **F1 Score** | **AUROC** |
| --- | --- | --- | --- | --- |
| BioClinicalBERT | 0.84 | 0.72 | 0.78 | 0.87 |
| BERT uncased | 0.49 | 0.63 | 0.55 | 0.74 |
| RoBERTa | 0.46 | 0.67 | 0.55 | 0.68 |
| XLNet | 0.77 | 0.65 | 0.71 | 0.87 |

AUROC = Area under the receiver operator characteristic.

BioClinicalBERT was selected as the final model for temporal validation because it had reasonable performance across all metrics (**Table 2**). XLNet and BioClinicalBERT achieved the same AUROC (0.87). RoBERTa achieved similar recall to XLNet but had low precision. BioClinicalBERT achieved an area under the precision-recall curve (AUPRC) of 0.73; A prediction probability threshold of 0.001842 was selected after maximizing recall and minimizing the false negative rate in training (**Figure 2**).

**Figure 2.**
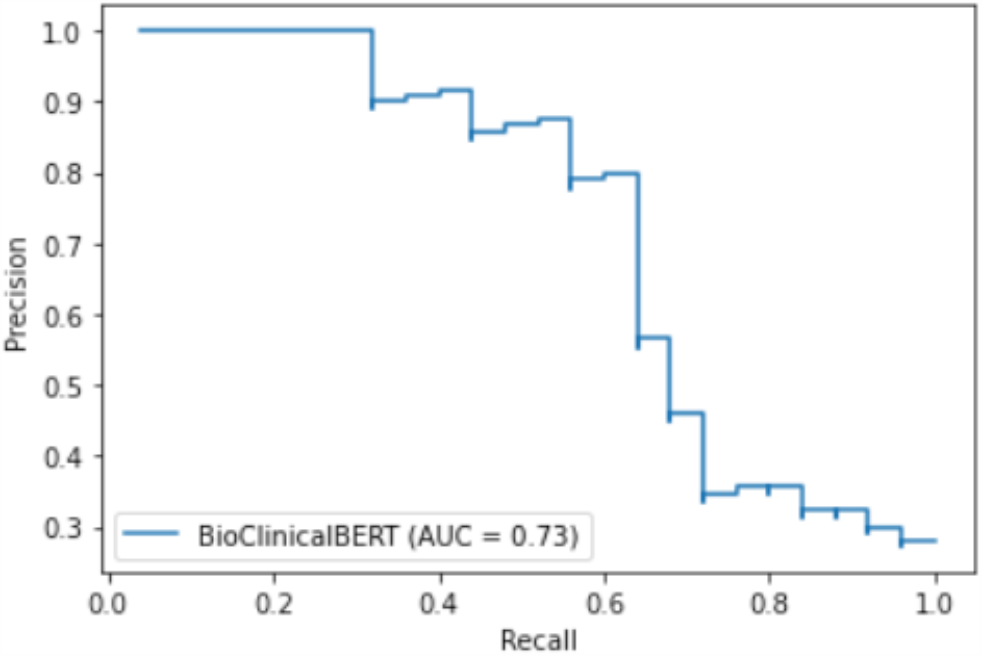
BioClinicalBERT precision-recall curve for threshold selection using the training set

### Temporal validation results

Temporal validation was conducted with the best performing model, BioClinicalBERT on all other patient notes following the first visit with an ADHD diagnosis (n=1020). After setting the threshold, 970 samples were classified as negative (not including a PTBM recommendation), and 50 samples were classified as positive. All 50 positively predicted notes and a random sample of 50 negatively predicted were examined in an error analysis.

BioClinicalBERT demonstrated improved performance in temporal validation (**Table 3**) with a recall of 0.92, representing low rates of false negative classifications (8%) and revealing that pediatricians recommended behavioral treatment in only 5% of non-first-ADHD visits.

**Table 3.** Best model (BioClinicalBERT) performance classifying clinical notes (annotated and unannotated)

| **Included notes** | **n** | **Recall** | **Precision** | **F1 Score** |
| --- | --- | --- | --- | --- |
| First ADHD visits | 127 | 0.72 | 0.84 | 0.78 |
| Other visits | 1020 | 0.92 | 0.70 | 0.80 |

### Error analysis

An error analysis was performed on predictions from the best performing model, BioClinicalBERT. The error analysis from temporal validation was conducted to understand when the model was misclassifying notes. The few false negative classifications missed mentions of in-office counseling on behavior management and mentions of names of specific therapists. A common false positive classification was recommendations for other types of behavioral treatment for co-occurring conditions documented in well-care visits (e.g., cognitive behavioral therapy for anxiety).

Most misclassified notes were cases with borderline prediction probabilities that were erroneously labeled as counseling on behavioral management. **Table 4** illustrates misclassification examples where note 1 is a false negative, and note 2 is a false positive. In note 1, a recommendation for behavior modification was provided, but it was describing a type of behavior modification without referral. In note 2, the false positive may have been caused by mention of a referral for developmental behavioral pediatrics (rather than a PTBM referral).

**Table 4.** Example notes with labels and predictions. Possible reasons for misclassification are **bold**, and true labels are also **<u>underlined</u>**.

| # | Example notes | Label | Prediction |
| --- | --- | --- | --- |
| 1 | well child care year old male icd cm. encounter for routine child health examination without abnormal findings z. plan hyperactivity likely adhd since present at school and at home. **<u>discussed setting rules and creating a schedule</u>** try to limit dyes and added sugars vanderbilt forms follow up with iep evaluation healthcare maintenance anticipatory guidance handout for age given. discussed growth nutrition dental care sleep activities exercise and safety issues. orders placed this encounter procedures vision test hearing screening test pure tone air only return in about year around. | 1 | 0 |
| 2 | female with the following diagnoses addressed today. screening for iron deficiency anemia poct hemoglobin. **behavior hyperactive external referral to development behavioral pediatrics** plan hgb is normal at.. encourage variety of foods. mom has scheduled a conference with teacher but meanwhile i will initiate a referral for developmental behavior peds to address concerns of inattentiveness and hyperactivity. f u as needed. | 0 | 1 |

## DISCUSSION

In this study, we demonstrated the feasibility of using an NLP algorithm (BERT) on a large and variable set of clinical notes to assess pediatrician adherence to guidelines in treatment of young children with ADHD. This approach allows to assess quality of care for the large number of children with ADHD (8-10% of pediatric population in the US), which would not be feasible by manual chart review. Leveraging NLP to assess quality-of-care is especially beneficial in assessing management of mental health disorders, where most data are descriptive and exist mainly in the unstructured text of the EHR, such as treatment recommendations that are not captured through electronic referrals. Our approach carries broad implications for using novel informatics approaches to assess quality of care, providing near real-time feedback to clinicians and health organizations, and driving quality improvement efforts that improve outcomes for individuals with mental and behavioral health disorders.

To the best of our knowledge, this is the first study to examine the use of pre-trained large language models (BERT) for ADHD quality of care measurement. An advantage to using BERT for classification is that models have already been trained on large datasets, meaning a large amount of data is not required to train models de novo. Despite the limited sample size in our study, we were able to fine-tune BERT and demonstrate reasonable performance in extracting adherence to guideline recommendations with our pipeline. Previous studies have examined using similar methods for information extraction for various use cases (e.g., medication management, patient monitoring)^27^; however, the application of these methods to assess quality of care and improve healthcare delivery is largely unexplored. In our work, we demonstrated the potential of using these methods for quality-of-care measurement.

The long-term goal of this work is to construct a quality officer and clinician-facing practice performance dashboard that would allow clinicians to understand how they are performing relative to evidence-based guidelines. For quality officers, the aim is to support them in auditing charts for quality-of-care assessments. Manual chart review is a time-intensive and painstaking process that can lead to delays in quality assurance (QA) processes. QA is a critical component of healthcare as it helps ensure that patients receive high-quality care. Currently, at the examined healthcare network, standard QA processes are conducted twice per year on a sample of patient notes, which does not facilitate actionable feedback to clinicians. Furthermore, the time-consuming task of manual chart review leads to limited assessment of care, focusing on narrow and accessible information in the chart (e.g., obtaining vitals in children prescribed ADHD medications). Such sub-optimal quality-of-care measurement, which represents the current status quo, are often discounted by health care organizations and clinicians as lacking in accuracy and clinical meaning.^28^ By automating the assessment of clinician adherence to established guidelines, the process can become more robust – assessing all aspects of evidence-based care for the entire population of children with ADHD rather than a small scope of care in a small sample, more efficient – reducing cost and time, more accurate - reducing human error and variability in the review process, and more actionable - allowing frequent reviews (e.g., monthly, quarterly) resulting in timely feedback to clinicians and healthcare organizations. An accurate and meaningful assessment of quality-of-care can lead to improved patient outcomes.

### Limitations

Although the study was conducted in a large network of primary care practices, which included a diverse population (including 14% Hispanic, similar to the US census), the sample size was limited, and the dataset was imbalanced with relatively few positive cases. To mitigate the imbalance, we leveraged BERT and evaluated the final model performance with metrics that are less sensitive to imbalanced classes such as precision and recall. In terms of generalizability, though the study was conducted in a single community-based pediatric network, the variable geographic locations of the practices (n=24) with varying available community services support the generalizability of the study. To further analyze generalizability, we plan to deploy the model on EHR data from other healthcare networks.

## CONCLUSION

Study findings suggest that implementing an NLP pipeline using a BERT model to capture treatment recommendations for children with ADHD offers a significantly better approach to quality measurement than the current status quo in healthcare organizations, which consists of limited claims-based metrics and/or laborious manual chart review of sample notes. Given the high prevalence of ADHD and the negative ramifications of inappropriate treatment recommendations commonly provided to young children with ADHD, the proposed approach offers a critical capability to address a large public health problem. This novel approach reduces data collection and reporting burden, assesses quality-of-care for an entire population of patients rather than a sample of patients, allows to measure multiple aspects of care, which are not captured in the current narrow-scope quality metrics, and enables near real-time feedback for clinicians and organizations to drive improvement efforts aimed at standardizing ADHD care and mitigating disparities. Ultimately, with expansion to additional quality metrics that combine structured and free-text (unstructured) data from the EHR, this NLP-powered approach will enhance clinician adherence to evidence-based practices and equitable treatment recommendations resulting in improved patient outcomes.

## Data Availability

All data produced in the present study are available upon reasonable request to the authors. The code used for this study is available via Github at https://github.com/ybannett/NLP_ADHD.

## Acknowledgments

We thank Packard Children’s Health Alliance and Stanford Research Information Technology for their support and assistance in data acquisition and extraction. We thank Dr. That Nam Tran (Sony) Ton, MD, for assistance in completion of chart review and annotation.

## Funding Source

Research reported in this publication was supported by the National Institute of Mental Health of the National Institutes of Health under Award Number K23MH128455 (Dr. Bannett). The content is solely the responsibility of the authors and does not necessarily represent the official views of the National Institutes of Health.

## Role of Funder

Funder did not have any part in design and conduct of the study; collection, management, analysis, and interpretation of the data; preparation, review, or approval of the manuscript; and decision to submit the manuscript for publication.

## Financial Disclosure

The authors have no financial relationships relevant to this article to disclose.

## Potential Conflicts of Interest

The authors have no conflicts of interest to disclose.

## Notes

### Competing Interest Statement

The authors have declared no competing interest.

### Author Declarations

This study was approved by the Stanford University School of Medicine institutional review board.

